# Identifying X-Chromosome Variants Associated with Age-Related Macular Degeneration

**DOI:** 10.1101/2023.08.28.23294688

**Authors:** Michelle Grunin, Robert P. Igo, Yeunjoo E Song, Susan H Blanton, Margaret A Pericak-Vance, Jonathan L. Haines, International Age-related Macular Degeneration Genomics Consortium

## Abstract

**Purpose:** In genome-wide association studies (GWAS), X chromosome (ChrX) variants are often not investigated. Sex-specific effects and ChrX-specific quality control (QC) are needed to examine these effects. Previous analyses identified 52 autosomal variants associated with age-related macular degeneration (AMD) via the International AMD Genomics Consortium (IAMDGC), but did not analyze ChrX. Therefore, our goal was to investigate ChrX variants for association with AMD.

**Methods:** We genotyped 29,629 non-Hispanic White (NHW) individuals (M/F:10,404/18,865; AMD12,087/14723) via a custom chip and imputed after ChrX-specific QC (XWAS 3.0) using the Michigan Imputation Server. Imputation generated 1,221,623 variants on ChrX. Age, informative PCs, and subphenotyeps were covariates for logistic association analyses with Fishers correction. Gene/pathway analyses were performed with VEGAS, GSEASNP, ICSNPathway, DAVID, and mirPath.

**Results:** Logistic association on NHW individuals with sex correction, identified variants in/near the genes *SLITRK4, ARHGAP6, FGF13* and *DMD* associated with AMD (P<1x10^-6^,Fishers combined-corrected). Via association testing of subphenotypes of choroidal neovascularization and geographic atrophy (GA), variants in *DMD* associated with GA (P<1x10^-6^, Fishers combined-corrected). Via gene-based analysis with VEGAS, several genes were associated with AMD (P<0.05, both truncated tail strength/truncated product P) including *SLITRK4* and *BHLHB9*. Pathway analysis using GSEASNP and DAVID showed genes associated with nervous system development (FDR: P:0.02), and blood coagulation (FDR: P:0.03). Variants in the region of a microRNA (miR) were associated with AMD (P<0.05, truncated tail strength/truncated product P). Via DIANA mirPath analysis, downstream targets of miRs show association with brain disorders and fatty acid elongation (P<0.05). A long-non coding RNA on ChrX near the *DMD* locus was also associated with AMD (P=4x10^-7^). Epistatic analysis (t-statistic) for a quantitative trait of AMD vs control including covariates found a suggestive association in the *XG* gene (P=2x10^-5).

**Conclusions:** Analysis of ChrX variants demonstrates association with AMD and these variants may be linked to novel pathways. Further analysis is needed to confirm results and to understand their biological significance and relationship with AMD development in worldwide populations.

## Introduction

Age-related macular degeneration (AMD) is the leading cause of blindness in the western world today for those over the age of 60 (Friedman 2004). The world’s aging population is due to double in the next 40 years^1–3^. Currently, the global cost of AMD is around $343 billion, with yearly direct costs from the United States medical system at $575 million dollars and $38,665 per person^4^. Current treatments for AMD focus on late stage symptoms, particularly the use of anti-VEGF antibodies to slow and control the growth of pathological new blood vessels through the choroid into the retina^5–7^.

The etiology of AMD is not fully understood, but age, smoking, and genetic variation, along with disorders such as hypertension, obesity, and diabetes, are contributing factors to this multifactorial condition^8–13^. The strongest known genetic risk factors include the *ARMS2/HTRA1* locus, as well as genes in the complement system, such as *CFH*^14^, *C2, C3, CFB*^15–17^, *CFI* and *C9*^18^ . Other genes and pathways are involved such as the *LIPC* gene in the lipid metabolism pathway, the VEGF pathway and receptor genes, and rare variants in multiple genes^19^. Currently, the largest dataset of AMD participants has been collected by the International Age-Related Macular Degeneration Genomics Consortium (IAMDGC), comprised of 24 centers worldwide who have collectively gathered 16,144 advanced AMD NHW participants and 17,832 NHW controls for genetic and phenotypic analysis, and discovered 52 novel and rare variants in 34 loci across the genome associated with AMD pathogenesis^19^. However, only ∼60 percent of the heritability of AMD is accounted for by the known risk variants, so investigation into the “missing heritability” of AMD will contribute greatly to the understanding of AMD pathogenesis.

Other studies have investigated transcription wide association (TWAS), protein expression in blood and retina, and analysis of microRNAs in AMD. However, the GWAS, TWAS, and other analyses performed did not include the entire genome, because the X-chromosome (X) was excluded. Unfortunately, only a small subset of the thousands of GWAS performed across different traits include X^20–22^, and most exclude X before analyzing, even if originally included in pre-quality control (PreQC) data^22,23^. Further studies that did include X sometimes use methods that did not account for the uniqueness of X^24–26^. This includes the obviously smaller male and female specific sample sizes, the impact of X inactivation in females vs males, reduced diversity on X, and sex-specific population structures, and of course association testing for autosomes incorrectly applied to X^20^. Fortunately, reanalysis and reinvestigation of X can be performed^27^. Specific quality control (QC) of X, such as sex stratified effects, clarifying the sample size of both men and women in the study, and the different variants between sexes on control samples can be done^21,26,28,29^.

X represents a significant portion (5%) of the nuclear genome, and 10% of the causes of Mendelian disorders^20^. Studies have highlighted the role of sexual dimorphism in many diseases such as autoimmune disorders, cancer, and psychiatric disorders. There is possible enrichment for sexually antagonistic alleles that can contribute to disease risk, and of course a difference between males and females on the genes that have unique functions on X^20,30–32^. X includes seven percent of the known microRNA (miRNA) and non-coding variants in the genome, which include long non-coding RNA (lincRNA) and others^33–35^. Approximately 16 percent of X-linked miRNA are implicated in immunity including those linked to the immune response and auto immune disease triggers. Dysregulation of the miRNA on X can contribute to some of these triggers^33–37^. Some autosomal miRNAs are associated with AMD. However, X-linked miRNAs have never been investigated^38–40^. Interaction between X linked miRNAs and genes located on and off X have been found in studies of multiple complex diseases^36^. Shared areas between intronic miRNA and known protein coding genes on X can lead to clusters working in tandem for biological function^36,37^.

Previous studies investigated 150 AREDS patients and 1,804 SNPs on X, and found a haplotype on X related with AMD^41^. This was a protective haplotype encompassing a 272 kb region, and covered the gene *DIAPH2*. The hypothesis at the time was an instability in X chromosome inactivation due to Primary Ovarian Failure, with a connection to *DIAPH2*^*42*^. There was one case report of a recombinant X mutation, causing a neurological phenotype and macular degeneration^43^. Winkler et al^44^ reanalyzed the IAMDGC dataset and tested for sex-stratified effects on the autosomes, without finding differences amongst the lead variants by sex, despite having 80 percent power in each sex to detect variants of interest. AMD is more frequent in women than men, and as such sex-specific analysis and chromosome-specific genetic analysis especially on large datasets are needed to fully unravel the genetic underpinnings of this disorder.

The IAMDGC GWAS^19^ used data imputed with the available reference panel at the time, but was unable to impute X due to software limitations. Imputing the European ancestry participants (EUR) with a more comprehensive reference panel, and including X, significantly increases the power of the IAMDGC dataset as a resource and allows for greater identification of variants contributing to AMD risk. The Haplotype Reference Consortium (HRC) contains 32,470 samples with 64,970 haplotypes of EUR ancestry, allowing extensive imputation improvement over the originally used 1000 Genomes Project. It also includes X, allowing analysis of X for the first time. We focused on the EUR ancestry to be consistent with the IAMDGC GWAS^19^.

Therefore, we set out to reanalyze the IAMDGC dataset, by first reimputing with the HRC to boost our power and coverage and then to analyze the X chromosome specifically to identify novel risk variants that could contribute to our understanding of AMD. We investigated X not only with methodology specific to the X-chromosome utilizing the new imputation, but also by multiple analyses including X-specific GWAS (XWAS) via variant and gene analysis, expression and eQTL analysis, non-coding RNA investigation including the microRNA and lncRNA that comprise so much of chromosome X, pathway and interaction analysis, and epistasis. This represents an exhaustive genetic analysis of the X chromosome in AMD.

## Methods

The IAMDGC SNP array data has ∼250K tagging and ∼250K rare/common variants, resulting in a starting point for pre-imputation of 569,645 variants genome wide. These were filtered according to previous methodology^19^. Briefly, only the individuals with a known phenotype (geographic atrophy (GA), neovascular AMD (nvAMD) as well as ophthalmalogically examined controls, along with NHW status based on the previous population stratification analysis^19^ were used in this analysis. Pre/post imputation pipelines were developed for an X specific GWAS, and analysis was performed with Plink 1.9 and XWAS 3.0, along with R and Bioconductor scripts. 18,865 female and 10,404 male IAMDGC samples were used (12,087 control/14,273 AMD), which included CNV/GA/Mixed as 4,916/1,482/942 (Table1). All data collection was approved by the institutional review board or ethics committee of all participating institutions.

**Table 1:**

|  | Female | Male |
| --- | --- | --- |
|  | 18,865 | 10,404 |
|  | Control | AMD |
| Phenotype | 12,087 | 14,273 |
| CNV: 4916 | GA: 1,482 | Mixed: 942 |

The Haplotype Reference Consortium panel (HRC) 1.1 dataset was used for the imputation reference panel. It contained 64,976 haplotypes including X. The Michigan Imputation Server (MIS) was utilized especially taking into account X-specific imputation. ShapeIT was used for pre-phasing, data on X was split by non-pseudoautosomal (non-PAR) and PAR regions. QC was performed via sex, for non-PAR and we excluded samples that did not match (n=56). Heterozygote genotypes were evaluated. 3 files were developed: the non-PAR female, PAR, and non-PAR male. Imputation was performed using minimac3.

The final variant count across the genome was 569,645 before imputation, but on chromosome X only 6,411 variants were present before imputation. The QC pre-imputation included a new identity by descent calculation (IBD), removal of the heterozygote haploid genotypes, Hardy-Weinberg equilibrium (HWE) calculated separately for males and females and the variants removed with a threshold of P<5.9x10^-8^, missing and frequency testing was performed according to standard (GENO > 0.1, MAF < 0.0005) and variants removed, along with removal of samples with missing phenotypes. 3 SNPs failed the sex frequency testing at a P=7.56x10^-6^, and were removed.

X was then imputed to 1,221,623 variants via the MIS and HRC 1.1. Post-imputation processing sensitive to sex occurred in similar steps to pre-imputation processing with similar cutoffs including HWE, missingness, frequency and sex-specific details, along with the QUAL threshold. There were still some SNPs with a different frequency amongst sexes in controls with P<5.9x10^-8^. After post-imputation processing the final number of variants on X was 597,585, and the final number of samples for the X analysis was 29,269 (M/F:10,404/18,865).

For analysis, informative PCs with threshold above eigenval 19 (PC 1,2,3,4,5,6,7), calculated with Plink 1.9 and considered informative; age, and sub-phenotypes were taken into account for all downstream analyses. The population structure on X captures a 1:2 male/female contribution, which is different than the autosomes. However, Chang^20^ evaluated both PCs taken from X and from autosomes, and determined that correction is more appropriate via the autosomes because of the small number of SNPs on X relative to the autosomes. Stouffers and Fishers correction were both used for analysis; results were similar.

Logistic regression was performed for a standard XWAS (X-wide association study), including covariates and sex-stratified. Logistic association was performed with sex correction, using both Fishers and Stouffers correction, including age and informative PCs as covariates. The significance threshold was set at 0.05/ the number of independent tests (pruned variants to account for linkage disequilibrium, final n = 164,976 variants) for a Bonferroni correction = 3.03x10^-7^. To look at X interaction, epistatic analysis was performed. The test used was a modified epistatic test that uses a t-statistic for a quantitative trait (XWAS 3.0).

VEGAS2 was utilized for gene based testing using both the truncated tail strength and the truncated product p-value testing. The suggestive P-value threshold was set at less than 0.05 for gene based testing, whereas the full significance value was set at 6.2x10^-5^ (genes in X chromosome n=804, 0.05/804 = 6.2x10^-5^). The threshold for significant SNPs to be included was set at 0.0001 for gene based analysis. We modeled males to be as equivalent to female homozygotes. In addition, we performed a sub-phenotype analysis, with GA alone, CNV alone, and a mixed analysis, where the mixed samples were added to GA and CNV separately. Using the XWAS suite, VEGAS2 was utilized for gene based testing using both the truncated tail strength and the truncated product p-value testing.

Pathway analysis programs were utilized to identify pathways of interest involving the most significant genes. Pathway analysis programs GSEASNP^45^ (http://www.nr.no/pages/samba/area_emr_smbi_gseasnp.) and ICSNPathway^46^ (http://icsnpathway.psych.ac.cn/) were used to test if a disease phenotype is influenced by genes enriched in a signaling pathway. GSEA-SNP modifies the original GSEA program which only took into account genes rather than SNPs of interest. We also utilized DAVID^47^, which takes into account genes that are found in clusters incorporating both GO and KEGG terms, as well as pathway and disease connections like OMIM (https://david.ncifcrf.gov/). Multiple programs were used to ascertain if genes/SNPs would overlap utilizing multiple methods, after performing pathway analysis.

Transcription factor motif analysis was performed using MEME, and the MEME suite of related programs (https://meme-suite.org/meme/)^48^.

## Results

### Single Variant and Gene-based Analyses

Using logistic regression with sex correction, variants in or near the genes *SLITRK4, ARHGAP6, FGF13* and *DMD* along with intergenic regions, were nominally associated with AMD (P<1x10^-6^; Fishers combined-corrected). (Table 2). all genes identified as significant (P<1x10^-7^) were found in genes expressed in eye, and may be involved in photoreceptor transduction or neurological disorders (Table2).

**Table 2:**

| Gene | Chr:BP | Fishers' corrected<br>value: variant | P- VEGAS<br>value: gene | P- SNPs per gene<br>(full set,<br>including those<br>in linkage) |
| --- | --- | --- | --- | --- |
| *SLITRK4* | X:142723657 | 2.5x10<sup>-7</sup> | 0.001 | 40 |
| *ARHGAP6* | X:11563246 | 1x10<sup>-6</sup> | P>0.05 | 2363 |
| *FGF13* | X:137852982 | 6.5x10<sup>-7</sup> | 0.03 | 3085 |
| *DMD* | X: 31217428 | 2x10<sup>-6</sup> | P>0.05 | 11712 |
| *MAGED1* | Gene based<br>only | P>1x10<sup>-6</sup> | 0.0004 | 202 |
| *BHLHB9* | Gene based<br>only | P>1x10<sup>-6</sup> | 0.0009 | 120 |

Pathway analysis using multiple programs (GSEASNP (GSEA4GWAS), DAVID, and ICSNPathway) after gene-based analysis on significant genes identified by VEGAS across all associated variants showed genes associated with nervous system development (FDR-P:0.02), and blood coagulation (FDR-P:0.03), both important pathways in AMD. These pathways have previously been identified as important with AMD, strengthening our results. In a paper published on IPA analysis, the nervous system, embryonic and organ development were the top three pathways found. (Zhang et al 2020 Journal of Ophthalmology), and these pathways were found by our analysis as well. In IPA analysis, pathways also identified variants in the blood coagulation pathway. The blood coagulation pathway has been found previously to be associated with response to VEGF in AMD patients^49^. DAVID pathway analysis of location based pathways (i.e., pathways that cluster in one tissue or location in the body, for example, eye, or blood cells) indicated clusters of significant genes located in brain or involved in locations involved in brain cancer. According to ICSNPathway analysis, the candidate causal SNP for the pathways identified was located in the gene GLUD2 (rs9697983) indicating the strength of that gene in the pathway found of electronic transport and metabolism (Table3).

**Table 3:**

| Pathway Analysis | Cluster Name | FDR |
| --- | --- | --- |
| DAVID | Cytoplasm | 0.04 |
| GSEA4GWAS | Endoplasmic reticulum network | 0.01 |
|  | Nervous System Development | 0.02 |
|  | Blood Coagulation | 0.03 |
|  | Wound Healing/Platelets | 0.03 |
| ICSNPathway | Electron Transport (GO:0006118) | 0.02 |
|  | Metabolism of Organic Acids, Amino Acids, Carboxylic Acids | 0.03 |
|  | (GO:0006520, GO:0019752). |  |

### Epistatic analyses

Every single variant not in linkage in the X chromosome was tested (n=164,976) for epistatic interactions. An suggestive epistatic interaction was found within X between a variant in the XG gene (X:2699555) and an intergenic variant (X:11868036), which was different between AMD patients and controls (P=2x10^-5^). This suggestive epistatic interaction was detected in the absence of main effects as neither of the locations had a main effect. Data from the Harvard Transcriptome project on expression of RNA from AMD patients and controls^50^ indicates that RNA in that area is expressed in AMD retina but not in control. According to the eye gTEX dataset^51^, the area between the genes *MSL3* and *FRMPD4* (near *ARHGAP6*), which contains the intergenic region found above via epistasic analysis, have some expression in retina, but it is not highly enriched (n=101 MGS1 retinas). XG was not mapped in that dataset at all, but would be between *ZBED1* and *CD99P1*, which both had some expression in retina but were not highly enriched.

### Subphenotype Gene Based Analyses

Sub-phenotype analysis found several genes with P<1x10^-6^, Fishers corrected. Analysis on CNV alone indicated an association with the gene *PHEX* (4916 cases/7926 controls). When including the mixed samples (CNV+mixed) an association was found with *ARHGAP6* and *PTCHD1*-AS (5828 cases/7926 controls). GA alone indicated an association with *DMD*, and one variant in *DACH2* (1482 cases/7926 controls). For the GA+mixed, all significant SNPs found were intronic (2424 cases/7926 controls). The results reaffirm the general X results and indicate a trend of genes that push the respective XWAS significant p-values amongst the sub-phenotypes. (SupplementaryTable1 for exonic hits)

### Non-Coding RNA Analyses

One long non-coding RNA (lncRNA), lnc-KDM6A1:2, was identified (4x10^-7^). Lnc-KDM6A1:2 is 717 bp, and functions as antisense. It has been validated by both the ENCODE and HAVANA datasets. According to the Harvard transcriptome it is present in normal human retina via SAGE data. A mirRNA, mir584, was associated in gene based testing, and is located near the *DMD* locus, which may be its target (RF0106, also known as AC090632.13/miRNA584-f), although the two loci are independent and not in linkage disequilibrium. This miRNA still needs to be validated, but is present in normal human retina according to the HARVARD transcriptome. All miRNA and lncRNA were checked via BLAST to confirm location and stem-loop sequence. It is part of the stringent set of the LNCpedia database, and can be found across Ensembl as well.

Several variants in the region of a microRNA(miR) were associated with AMD (VEGAS2, P<0.05, truncated tail strength and truncated product P, Table 4). Only the variants with the strongest association signals (P<1x10^-6^) are included in the gene-based test, allowing us to use a lower p-value for gene based associations. The locations of these miRNA are near *PHEX* and *DMD*. These miRNAs may directly influence nearby genes. MiRNA were scanned by both the 3p and the 5p to determine their sequence and location. Via ENCODE data, these miRNAs were near some regulatory elements that regulate areas in the cerebrum and embryonic cell development (location: X:1427222).

**Table 4:**

| miRNA | Chr:BP | P-value |
| --- | --- | --- |
| miR-220 | X:122695945-122696055 | 0.03 |
| miR-652 | X:10939960-10940057 | 0.003 |
| miR-548 | X:22191615-22191699 | 0.001 |
| miR-584 | X:31356817-31356938 | 0.04 |
| miR-934 | X:135633036-135633119 | 0.004 |
| miR-2114 | X:149396238-149369318 | 0.04 |
| miR-509-1 | X:146340277-146341244 | 0.05 |

To determine the predicted function for the miRNAs initially identified via VEGAS2, DIANA mirPath analysis was utilized to identify downstream targets of these miRs using KEGG pathways (Table 5). Results show association with brain disorders and fatty acid elongation (P<0.05), particularly for Prion diseases (gene targets *MAP2K2* and *PRNP*) and Huntington’s disease (gene targets *HTT, CLTA, NDUFS1, COX4I1, EP300, SOD2, TBP*, and *UQCRB*). .

**Table 5:**
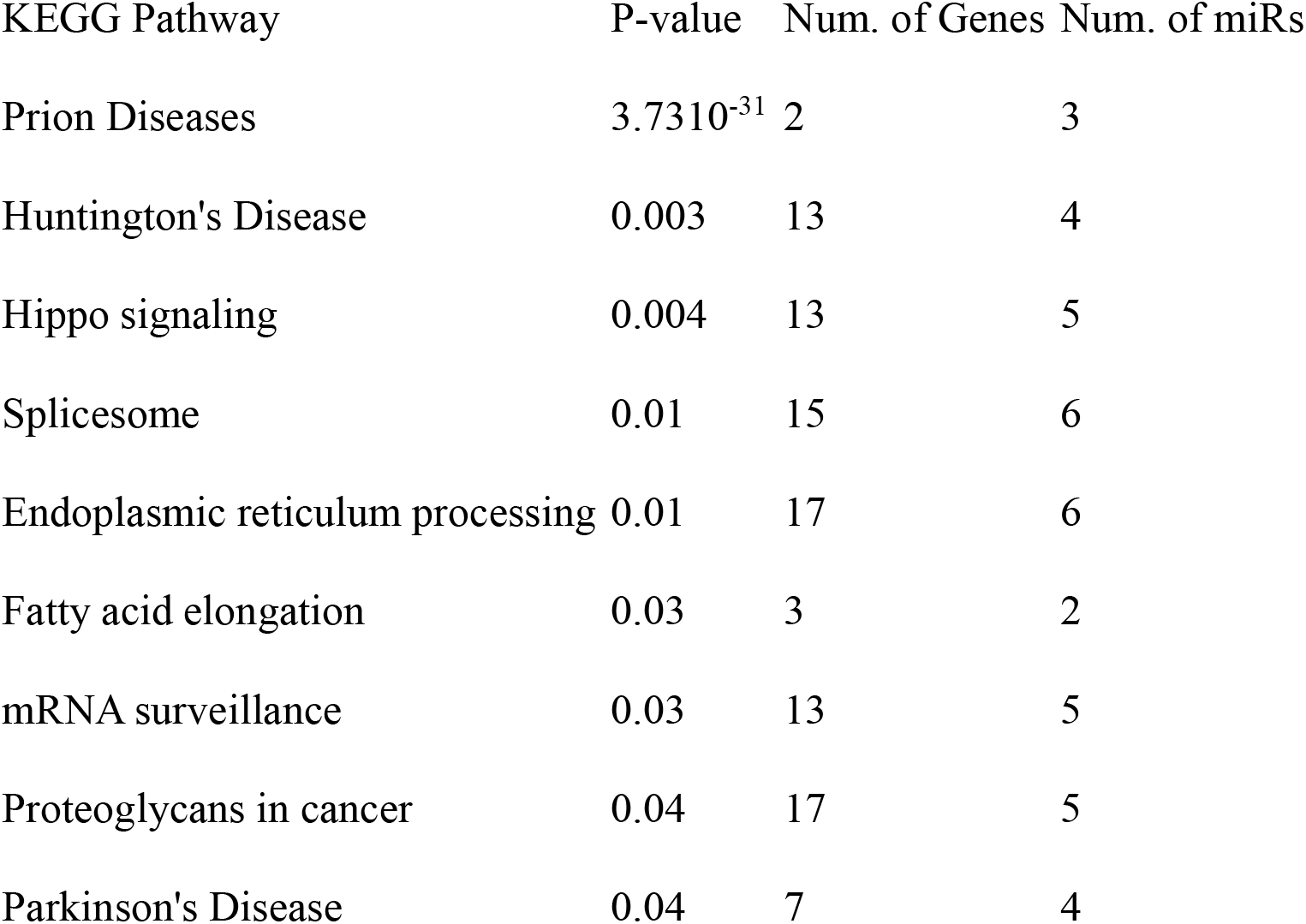

Transcription factor motif analysis was performed using the MEME suite of programs. Motifs were investigated in genes that were associated with AMD via VEGAS gene-based testing. This was performed with an unbiased analysis, one or more motifs found in sequences, and a maximum of 5 motifs per sequence with P<1x10^-7^. DREME motif analysis for short motifs was also performed to investigate an e-value < 0.01 (considered significant as the significance threshold for motifs is E = 0.05), but no results were found. GOMO analysis on promoters was performed on GO terms where the q-values were <0.05. Once motifs were identified, the HOCOMOCO database for human-specific motifs associated with transcription factors was utilized with TOMTOM to identify motifs that were associated with transcription factors found in multiple genes associated with AMD. This was compared to random with over 10 permutations to clarify the null hypothesis. An association to a motif found in multiple sequences of AMD was found for the transcription factors IRF3 (interferon), PRDM6 (involved in histone activity), and ZFP82 (involved in DNA binding and zinc-finger activity), along with others of lesser significance. (Table 6)

**Table 6:**

| Transcription<br>Factor | P-value | E-value | Motif<br>Overlap |
| --- | --- | --- | --- |
| IRF3 | $9.59 \times 10^{-7}$ | $3.86 \times 10^{-4}$ | 20 |
| PRDM6 | $3.12 \times 10^{-6}$ | $1.25 \times 10^{-3}$ | 13 |
| ZFP82 | $9.02 \times 10^{-6}$ | $3.63 \times 10^{-3}$ | 24 |
| IRF1 | $1.55 \times 10^{-5}$ | $6.25 \times 10^{-3}$ | 20 |
| BC11A | $1.73 \times 10^{-5}$ | $6.97 \times 10^{-3}$ | 17 |
| NFAC1 | $2.18 \times 10^{-5}$ | $8.75 \times 10^{-3}$ | 15 |
| SPIB | $5.01 \times 10^{-5}$ | $2.02 \times 10^{-2}$ | 17 |
| SPI1 | $7.21 \times 10^{-5}$ | $2.90 \times 10^{-2}$ | 17 |
| IRF2 | $7.75 \times 10^{-5}$ | $3.11 \times 10^{-2}$ | 20 |
| STAT2 | $8.67 \times 10^{-5}$ | $3.49 \times 10^{-2}$ | 19 |
| IRF8 | $9.38 \times 10^{-5}$ | $3.77 \times 10^{-2}$ | 20 |
| ZN394 | $2.10 \times 10^{-4}$ | $8.43 \times 10^{-2}$ | 20 |
| GATA3 | $2.60 \times 10^{-4}$ | $1.05 \times 10^{-1}$ | 11 |
| ETS2 | $2.93 \times 10^{-4}$ | $1.18 \times 10^{-1}$ | 13 |
| SOX2 | $3.43 \times 10^{-4}$ | $1.38 \times 10^{-1}$ | 13 |
| FLI1 | $4.55 \times 10^{-4}$ | $1.83 \times 10^{-1}$ | 18 |
| ANDR | $6.56 \times 10^{-4}$ | $2.64 \times 10^{-1}$ | 18 |

## Discussion

In previous analyses of AMD, the X chromosome was not included despite its many known roles in human biology. The Y chromosome and its association with age and AMD has already been studied, but no study until now has looked at X^52^. We hypothesized that X chromosome variants are associated with AMD pathogenesis, and that these variants may be linked to novel pathways. We identified multiple novel loci that associate with AMD status on the X chromosome, including some found in the loci involved in the non-coding genome. Therefore, although the X chromosome has been consistently overlooked in GWAS, its importance cannot be. AMD pathology is related to the regulation of the classical and non-classical immune systems, through the non-coding genome. We further explored the potential role of miRNAs and epistatic interactions between loci on the X chromosome and other loci that may contribute to AMD that have not yet been identified. The genes identified include some that are associated with other retinal disorders, or are regulators of other pathways like the non-coding RNA identified. No analysis has been done previously on the pathways that significantly involve X, nor on the non-coding RNA that has been found here to understand their role in AMD, and it allows the greater understanding of AMD heritability to be able to be understood from a full look at the X chromosome.

Pathways identified include both neuronal and eye pathways, as well as lesser studied pathways like wound healing and platelet function^53^These analyses, both gene and non-coding RNA based, as well as incorporating both pathway and epistatic interactions, provide a novel and important piece in the genetic puzzle of AMD that has not been examined until now. Many of these genes may represent novel targets, and the interactions of these miRNA and lncRNA with the genes on the X chromosome, as well as other genes across the genome has not been studied fully.

The genes identified were all expressed in retina. *DMD* is the protein for dystrophin, mutations in which cause Duchenne muscular dystrophy (DMD); in the retina it actually is involved in signaling and is located in photoreceptor terminals, retinal neurons, cone/ rod synapses, and other parts of the photoreceptor synaptic complexes^54^. It is involved in the optical neuron-bipolar pathways as well as the plasma membrane in Muller cells^55^. Most patients with DMD have some abnormal ERG results, including ‘negative’ scotopic ERGs, and others, especially those with deletion of the gene. There are also differential isoforms based on the various mutations/deletions, which associate with phenotype. This indicates that dystrophin continues to play a role in phototransduction^54,56^. In addition, *HTRA1*, one of the major genetic risk factors for AMD, is upregulated in DMD and may play a role in cell growth, influencing abnormal growth in the disease or affected cell growth or tolerance for repair.

*ARHGAP6* is a rhoGAP family member. The activation of the enzyme GTPAse protein with a specificity for rhoA and a cytoskeletal protein for actin remodeling. *SLITRK4* is an axonal growth controlling protein. *FGF13* is a fibroblast growth factor, which is involved in mitogenic and cell survival, tissue repair, morphogenesis and cell growth. All of these genes were expressed in retina according to the Harvard Retinal Transcriptome, and could be novel targets for either drug therapy or in how they regulate the other AMD known loci.

There was an association found with miRNA targeting genes involved in prion diseases in the pathway analysis (hsa05020). Prion diseases have a subset of “pathogenic” non-coding RNAs that are involved in the disorder, and these same miRNA are involved in AMD and Alzheimer disease (AD)^57,58^. In addition, the protein aggregation process in AMD and AD have a similarity to prion disease, and some classify them as “prion-like” diseases. All prion-like diseases cause retinal damage in humans, like the plaques of amelyoid-beta found in AD, but also in AMD^59^, and therefore they can be thought of as related to each other, not just in prion disease itself which has its own photoreceptor degradation, but in the pathogenesis involved in these diseases^60^.

This study represents the first study performed on the full X chromosome in AMD and includes a novel lncRNA association as well as genomic variants nominally associated with AMD pathogenesis. Further investigation is needed to clarify the roles of these identified genes and loci. This could involve via silencing/knockdown or overexpression of miRNA/lncRNA in RPE cell lines to evaluate function and survival^38,61,62^, or utilizing the MERFISH technique for localization in situ inside the cell for postranscriptional regulation^63,64^. Further analysis is needed to confirm these results and to understand their biological significance and relationship with development of AMD in worldwide populations.

Michelle Grunin^1,2^, Robert P. Igo, Jr^1,2^, Yeunjoo Song^1,2^, Susan Blanton^3^, Margaret Pericak-Vance^4^, Jonathan L. Haines^1,2^, International Age-related Macular Degeneration Genomics Consortium

## Author Contributions

MG, RPIJr, YS analyzed data. MG and JLH drafted and critically reviewed the manuscript. YS, SB, MP-V, JLH developed the ideas, advised on results, and critically reviewed the manuscript.

## Data availability statement

The genotype data analyzed during the current study were generated by the IAMDGC and are available through the database of Genotypes and Phenotypes (dbGAP; Accession: phs001039.v1.p1). Summary statistics for the IAMDGC data is available currently at http://amdgenetics.org.

## Ethical approval

The IRB of Case Western Reserve University -University Hospitals (IRB Number EM-14-04) gave ethical approval for this work.

The study participants were previously ascertained by IAMDGC cohorts as described in Fritsche et al, 2016, Nature Genetics. All participants provided informed consent, and the study was approved by institutional review boards as previously described.

